# Evidence of a significant secretory-IgA-dominant SARS-CoV-2 immune response in human milk following recovery from COVID-19

**DOI:** 10.1101/2020.05.04.20089995

**Authors:** Alisa Fox, Jessica Marino, Fatima Amanat, Florian Krammer, Jennifer Hahn-Holbrook, Susan Zolla-Pazner, Rebecca L. Powell

**Affiliations:** Department of Medicine, Division of Infectious Diseases, Icahn School of Medicine at Mount Sinai, New York, NY, USA; Department of Psychology, University of California Merced, Merced, CA, USA; Department of Microbiology, Icahn School of Medicine at Mount Sinai, New York, NY, USA

## Abstract

SARS-CoV-2, commonly termed COVID-19 for the illness it causes, has infected >3.2 million people, including >220,000 deaths. Human milk IgG originates mainly from blood, therefore a SARS-CoV-2-reactive antibody (Ab) response in milk would be expected (1). However, IgG comprises only ~2% of milk Ab, with most milk Abs originating from mucosa-associated lymphatic tissue (1). Therefore, the extent of the milk immune response to SARS-CoV-2 is unknown (2). This response is critical for infants and young children, who tend not to suffer greatly from COVID-19 pathology but are likely responsible for significant virus transmission (3-5). Perhaps even more significant is the fact that milk Abs could be purified and used as a COVID-19 therapeutic, given they would likely be of the secretory (s) class and highly resistant to proteolytic degradation in the respiratory tissue (2, 6). In this preliminary report, 15 milk samples obtained from donors previously-infected with SARS-CoV-2 as well as 10 negative control samples obtained prior to December 2019 were tested for reactivity to the Receptor Binding Domain (RBD) of the SARS-CoV-2 Spike protein by ELISAs measuring IgA, IgG, IgM, and secretory Ab. Eighty percent of samples obtained post-COVID-19 exhibited IgA reactivity, and all these samples were also positive for secretory Ab reactivity, suggesting the IgA is predominantly sIgA. COVID-19 group mean OD values of undiluted milk were significantly greater for IgA (p<0.0001), secretory-type Abs (p<0.0001), and IgG (p=0.017), but not for IgM, compared to pre-pandemic group mean values. Overall, these data indicate that there is strong sIgA-dominant SARS-CoV-2 immune response in human milk after infection in the majority of individuals, and that a comprehensive study of this response is highly warranted.

## Background

In December 2019 several cases of atypical pneumonia were reported in Wuhan, China. It was soon determined that this illness, now named COVID-19, was due to a newly identified coronavirus, termed SARS-CoV-2 (7). On March 11, 2020, the WHO declared the SARS-CoV-2 outbreak a pandemic. To date, SARS-CoV-2 has infected over 3.2 million people worldwide, including >220,000 deaths. Though COVID-19 pathology in children is typically mild compared to adults, approximately 10% of infants experience severe COVID-19 illness requiring advanced care (8, 9). Importantly, as COVID-19 pathology does not correlate with transmissibility, infants and young children are likely responsible for a significant amount of SARS-CoV-2 dissemination (3–5); therefore, protecting this population from infection remains essential. One potential mechanism of protection might be passive immunity via breastfeeding by a previously-infected mother. *If this response is potent, this is also relevant as a potential COVID-19 therapeutic*. These milk antibodies (Abs) may be far more effective than convalescent plasma or immunoglobulin.

As the pandemic continues, many pregnant and breastfeeding women will be infected with SARS-CoV-2. To date, no evidence of vertical transmission in utero has been found. Yet, sporadic cases of newborns infected with SARS-CoV-2 have been reported, with transmission most likely occurring during delivery (10–12). In terms of transfer of SARS-CoV-2 immunity to the neonate, data from the SARS (SARS-CoV-1) outbreak may give insight, but are conflicting: in one case, a woman was infected in the 7^th^ week of pregnancy. At near-term delivery, her serum was positive for SARS antibodies (Abs) but cord blood and placenta were negative. Milk samples were also negative (13). In another case, a woman infected in the 19th week of pregnancy was positive for SARS Abs in serum, cord blood, and milk shortly after delivery (14). One larger study of 12 pregnant women infected with SARS found that all babies’ serum were negative for SARS Ab after birth (15). As such, placental transfer of maternal Ab against SARS-CoV-2 may not necessarily occur, which suggests young infants are at particular risk for infection. Furthermore, infants of mothers infected post-partum would certainly depend on milk Ab for protection.

As milk IgG originates predominantly from serum, it follows that specific IgG in milk should appear contemporaneously with the serum Ab response, though IgG comprises only ~2% of milk Ig (2). Approximately 90% of human milk Ab is IgA and ~8% IgM, nearly all in secretory (s) form (sIgA/sIgM; polymeric antibodies (Abs) complexed to j-chain and secretory component (SC) proteins) (1, 2, 16). Nearly all sIgA/sIgM derives from the gut-associated lymphoid tissue (GALT), though there is also homing of B cells from other mucosa (i.e. the respiratory system) to the mammary gland; therefore, we should expect some SARS-CoV-2-specific sIgA/sIgM to be present in milk (1). Importantly, *in vivo* studies have demonstrated that secretory Abs are durable, transit well to the mucosa after systemic administration, and are resistant to enzymatic degradation; therefore, should SARS-CoV-2-reactive Abs be produced at high titer after infection, these predominantly secretory-type Abs have great potential as a COVID-19 therapeutic (2, 6).

To date, the magnitude, functionality, and durability of the human milk immune response to SARS-CoV-2 is currently unknown. Therefore, SARS-CoV-2-reactive milk Abs must be comprehensively studied for their potential therapeutic and protective efficacy. Towards that aim, we have recruited over 1000 lactating participants, including over 350 who have recovered from COVID-19 illness. This report details the preliminary findings regarding SARS-CoV-2-reactive IgA, IgG, IgM, and total secretory-type Ab in the milk of 15 donors from this cohort.

## Materials and Methods

### Human Milk

Approximately 30mL of milk was obtained from consented study participants using electronic or manual pumps. Each participant was recruited and interviewed in accordance with an IRB-approved protocol. Participants either had a laboratory-confirmed COVID-19 infection, or highly likely infection based on close contact with a confirmed COVID-19 case and/or symptoms of infection such as cough, anosmia, malaise, diarrhea, and fever. Milk was obtained ~14-30 days after symptoms had abated. Milk was pumped by the participants and frozen in their homes until sample pickup. Milk samples were then thawed, centrifuged at 800g for 15 min, fat was removed, and supernatant transferred to a new tube. Centrifugation was repeated 2x to ensure removal of all cells and fat. Skimmed acellular milk was aliquoted and frozen at −80C until use. Pre-pandemic negative control milk samples were obtained in accordance with IRB-approved protocols prior to December 2019 for other studies, and had been stored in laboratory freezers at −80C before processing exactly as described for COVID-19 milk samples.

### SARS-CoV-2 ELISA

A SARS-CoV-2 ELISA using blood plasma was recently developed and validated and we have adapted this assay for use with human milk (17, 18). Briefly, half-area 96-well plates were coated with the Receptor Binding Domain of the SARS-CoV-2 Spike protein, produced recombinantly as described (18). Plates were incubated at 4C overnight, washed in 0.1% tween/PBS (PBS-T), and blocked in PBS/3% goat serum/0.5% milk powder/3.5% PBS-T for 1h at room temperature. Milk was diluted in 1% BSA/PBS and added to the plate. After 2h incubation at room temperature, plates were washed and incubated for 1h at room temperature with horseradish peroxidase-conjugated goat anti-human-IgA, goat anti-human-IgM, goat antihuman-IgG (Rockland), or goat anti-human-secretory component (MuBio) diluted in 1% BSA/PBS. Plates were developed with TMB reagent followed by HCl and read at 450nm on an ELISA plate reader.

## Results

### Most milk (80%) obtained post-COVID-19-recovery exhibits IgA reactivity to RBD

Fifteen milk samples obtained from donors previously-infected with SARS-CoV-2 as well as 10 samples obtained as part of other studies prior to December 2019 were tested in duplicate in 3 unique experiments for separate assays measuring IgA, IgG, IgM, and secretory-type Ab reactivity (the secondary Ab used in this assay is specific for free and bound SC). The 10 pre-pandemic control undiluted milk samples were used to determine positive cutoff values for each assay, calculated as the mean OD + 2^*^SD. These control samples exhibited some low-level reactivity in all 4 assays; however, this non-specific or cross-reactive binding was notably greater for the IgA assay. Despite the higher positive cutoff value for this assay, 12/15 milk samples obtained from previously-COVID-19-infected donors exhibited IgA reactivity to RBD significantly above this cutoff value when undiluted, with wide variation in the ultimate binding endpoint (Fig. 1a, first panel). All 12 of these samples were also positive for secretory-type Ab reactivity to RBD (Fig. 1a, second panel). The IgA and secretory Ab (SC) OD values for undiluted milk were found to be highly correlated (r=0.81, p<0.0001 by Spearman correlation test; Fig. 2).

**Figure 1.**
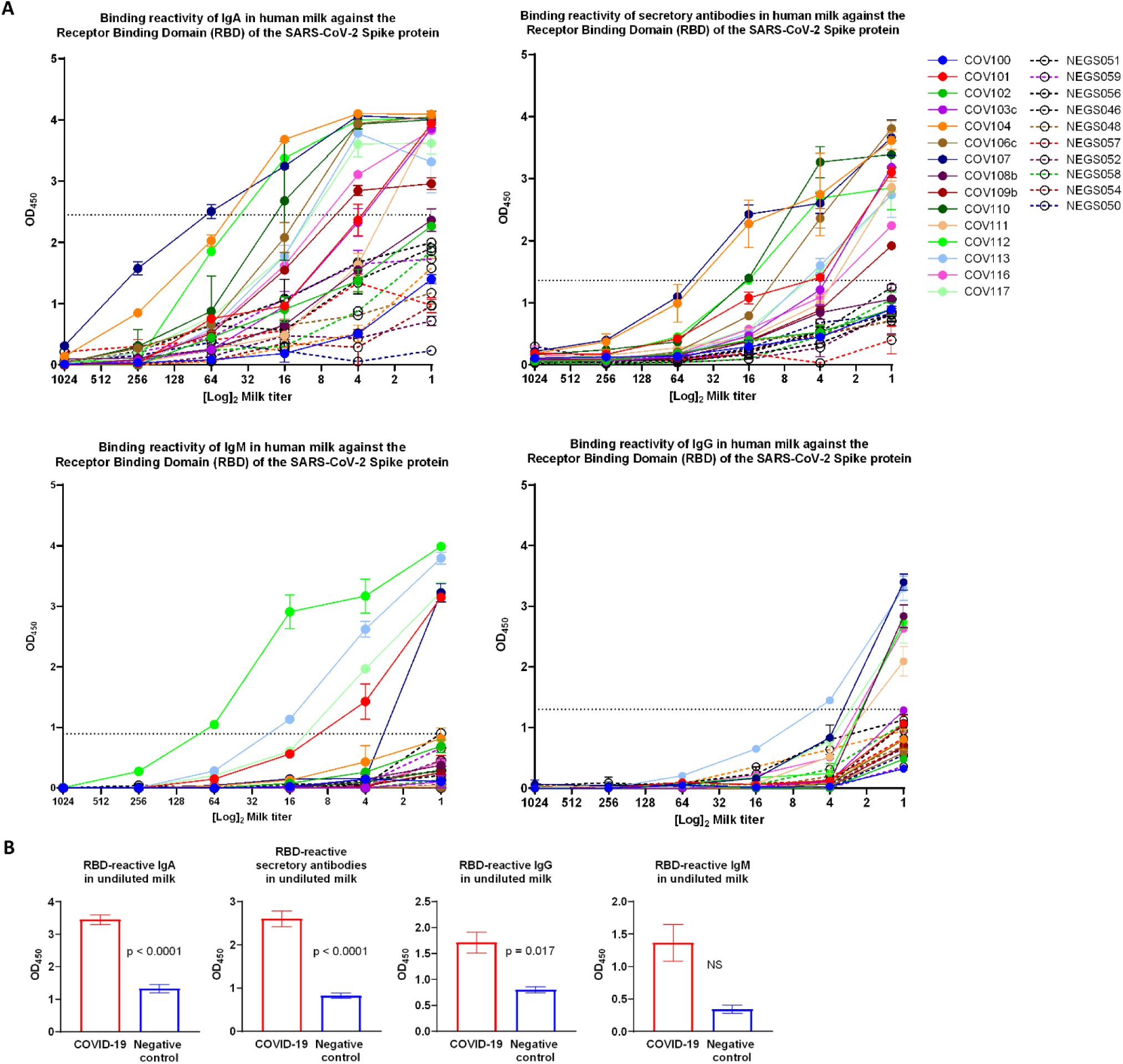
Distinct reactivity by ELISA of human milk against SARS-CoV-2 among samples collected from previously-infected donors. Briefly, plates were coated with the receptor binding domain (RBD) of the SARS-CoV-2 Spike protein overnight. Plates were washed, blocked, and titrated human milk obtained from 10 donors before December 2019 (NEG; segmented lines) and from 15 donors recovered from previous COVID-19 infection (COV; solid lines) was added to the plate. After incubation, plates were washed and incubated with the appropriate secondary antibody and read on an ELISA plate reader. Samples were run in duplicate. This experiment is representative of 3 unique repeats. (A) shows the full titration data with positive cutoff value (dotted line) calculated as the mean OD for undiluted milk + 2*SD. (B) shows grouped OD values for undiluted milk. Mean with SEM is shown for all data. Groups were compared by 2-tailed Mann-Whitney test. NS: not significant.

**Figure 2.**
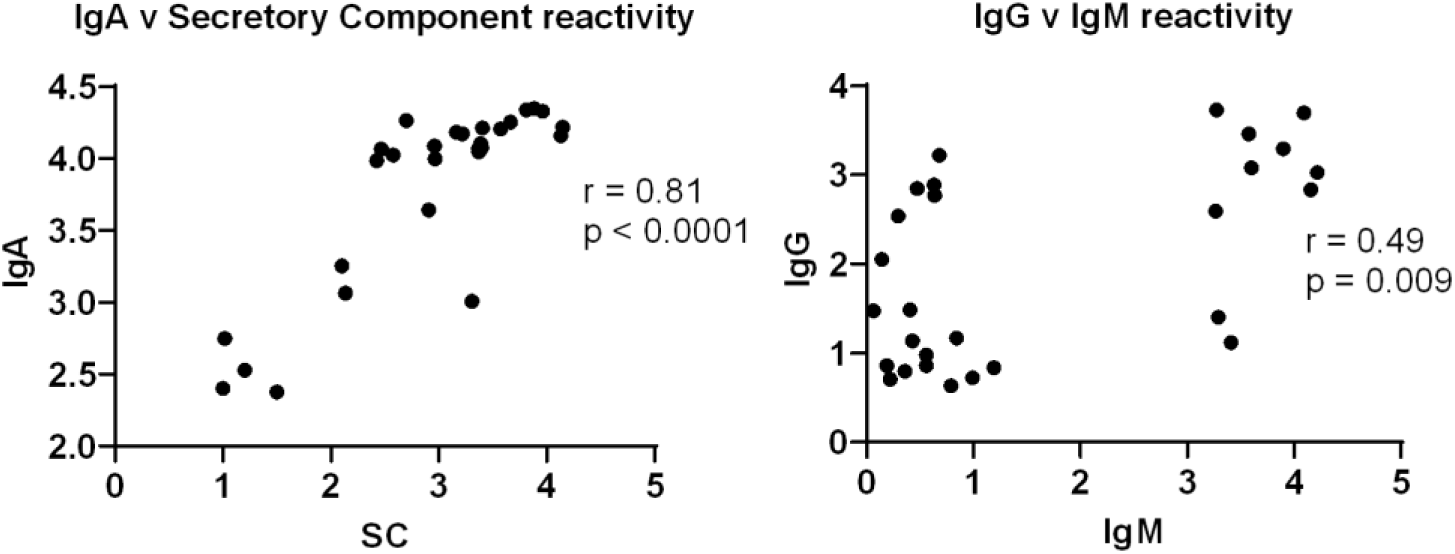
Correlated IgA/secretory antibody and IgG/IgM reactivity. OD values for undiluted milk were used in duplicate in a 2-tailed Spearman correlation test. SC: secretory component-containing antibodies.

### The RBD-reactive IgA response appears dominant and is not necessarily concurrent with a measurable IgG or IgM response

Of the 12 milk samples shown to be positive for IgA/SC reactivity, 4 samples also exhibited positive IgG and IgM reactivity to RBD (Fig. 1a). An additional 2 samples exhibited positive IgG reactivity but not IgM, and 1 sample also exhibited IgM reactivity but not IgG. One sample exhibited only positive IgG reactivity, but did not exhibited any IgA, IgM, or SC reactivity above the positive cutoff. These data are summarized in Table 1.

**Table 1.** Sample reactivity profiles by subclass.

| Sample ID | IgA | SC | IgG | IgM |
| --- | --- | --- | --- | --- |
| COV107 | + | + | + | + |
| COV112 | + | + | + | + |
| COV113 | + | + | + | + |
| COV117 | + | + | + | + |
| COV111 | + | + | + | - |
| COV116 | + | + | + | - |
| COV101 | + | + | - | + |
| COV103c | + | + | - | - |
| COV104 | + | + | - | - |
| COV106c | + | + | - | - |
| COV109b | + | + | - | - |
| COV110 | + | + | - | - |
| COV108b | - | - | + | - |
| COV100 | - | - | - | - |
| COV102 | - | - | - | - |

Overall, OD values of undiluted milk obtained from COVID-19-recovered donors and pre-pandemic controls for each assay were grouped and compared, and it was found that the COVID-19-recovered group mean values were significantly greater for IgA (p<0.0001), secretory-type Abs (p<0.0001), and IgG (p=0.017), but not for IgM (all groups were compared by 2-tailed Mann-Whitney test). As well, OD values for undiluted milk were compared between each assay (IgA v IgG, IgA v IgM, IgM v SC, IgM v IgG, and IgM v SC). It was found that no correlation existed between the reactivities of these Ab subclasses, with the exception of IgM v IgG, which were found to correlate (r=0.49, p=0.009; Fig. 2).

## Discussion

The data presented herein is preliminary using a small sample size and only serves to suggest what might be the typical range of the antibodies generated in human milk following SARS-CoV-2 infection. The samples analyzed represent only a snapshot of what is likely a dynamic immune response. A much larger sample size and long-term follow-up study is needed to better understand SARS-CoV-2 immunity in milk, as well as whether a typical response is truly protective for breastfed babies or if this response would generate sufficient Abs to be purified and used therapeutically to treat COVID-19 illness.

Though it might be expected that the milk Ab response would be reflective of systemic immunity – i.e., milk Ab should generally mirror serum Ab – only a small fraction of milk Ab originates from serum – likely less than 10%, and only ~2% of milk Ab is IgG (2). Human milk Ab is ~90% IgA and 8% IgM, nearly all sIgA/sIgM. These polymeric Abs are complexed to SC, which is the cleavage product of the pIgR that marks these secretory-type Abs as it transports them into the milk (1). SC is essential for the survival of secretory Abs in relatively harsh mucosal environments such as the infant gut where they would otherwise succumb quickly to proteolytic degradation (1). The B cells that ultimately produce milk sIgA/sIgM originate mainly from the GALT, known as the *entero-mammary link*, with some proportion originating from other mucosa such as the respiratory system (1, 19, 20). Evidence supporting this *entero-mammary link* include the finding that mammary gland B cells display an adhesion receptor profile matching that found on GALT B cells, and that milk sIgA exhibits high reactivity for enteric antigens compared to blood (21–24). In fact, PRM/Alf mice (possessing elongated intestinal tracts) have been shown to exhibit increased gut-derived B cells in the mammary gland and increased milk sIgA concentration compared to wild type mice (25). As such, there is much precedent for milk Ab composition and specificity being unique from that found in blood, as the data presented here suggests. Though we did not compare the milk donors’ blood Ab titers to the milk data, it was evident that most of the samples contained SARS-CoV-2-reactive IgA without necessarily containing measurable IgG and/or IgM, which particularly in the case of IgG, would likely be derived in large part from the serum. Notably, IgG and IgM reactivities in undiluted milk exhibited a moderate correlation. It may also be that as total IgG and IgM are so much lower in milk than IgA, that this ELISA lacked the sensitivity to pick up low-titer responses. Notably, one sample, COV108b, exhibited only IgG reactivity. Future experiments will compare milk and serum Ab titers.

Though it has been determined by previous studies that most IgA in human milk is sIgA, our ELISA could not determine with certainty that the IgA (or IgM) measured was of the secretory type or not (1). The assay measuring secretory Ab reactivity employs a secondary Ab specific for the SC, which can be free, or bound to Ab. Notable, all samples exhibiting positive IgA reactivity also exhibited positive SC reactivity, and a very strong positive correlation was present when comparing the OD values of undiluted milk for the IgA and SC assays. This suggests that a very high proportion of the SARS-CoV-2-reactive IgA measured herein was sIgA. This is extremely relevant to the possibility of using purified milk Ab as a COVID-19 therapy, as this sIgA-dominant milk Ab is highly unique from the IgG-dominant convalescent plasma or purified plasma immunoglobulin being tested currently. This sIgA-rich therapeutic would likely survive well upon targeted respiratory administration, where a much lower dose of Ab might be needed for efficacy compared to that used systemically. Moreover, non-human primate studies have found that dimeric IgA given systemically transits well to mucosal compartments, and maintains high levels in blood for a significantly longer period after administration compared to IgG or monomeric IgA (6).

As we continue with our comprehensive studies on the human milk immune response to SARS-CoV-2, we aim to ultimately determine the efficacy of ‘convalescent milk Ab’ as a treatment for COVID-19, and the utility of these Abs to prevent or mitigate infant SARS-CoV-2 infection. These data will have implications beyond the pandemic, as they will serve to fill relatively large knowledge gaps regarding human milk immunology. As well, as SARS-CoV-2 vaccines are developed, these data will provide a comparative baseline that will be critical for understanding the efficacy of vaccines in terms of the passive immunity they generate to protect breastfed babies.

## Data Availability

The authors confirm that the data supporting the findings of this study are available within the article [and/or] its supplementary materials.

## Acknowledgements

As always, we are indebted to the milk donors who make this work possible.

